# A Simple Electronic Medical Record System Designed for Research

**DOI:** 10.1101/2020.10.25.20219287

**Authors:** Andrew J King, Luca Calzoni, Mohammadamin Tajgardoon, Gregory F Cooper, Gilles Clermont, Harry Hochheiser, Shyam Visweswaran

## Abstract

With extensive deployment of electronic medical record (EMR) systems, EMR usability remains a major source of frustration to clinicians. There is a significant need for a simple EMR software package that will enable investigators to study design and usability in a laboratory setting. We developed an open-source software package that implements the display functions of an EMR system. The user interface emphasizes the temporal display of data such as vital signs, medication administrations, and laboratory test results. It is well suited to support research about clinician information-seeking behaviors and adaptive user interfaces in terms of measures that include task accuracy, time to completion, and cognitive load. The Simple EMR System is freely available to the research community on GitHub.

## 1 Background

The user interfaces of electronic medical record (EMR) systems are a hodgepodge of legacy designs, billing requirements, and local customizations. The resulting misalignment of clinical and electronic workflow adds extraneous work and is a major source of clinician frustration [1]. To remedy the ills of the EMR, greater attention needs placed on understanding EMR use, design consequences [2], and potential improvements. Unfortunately, laboratory-based research of clinician-EMR interactions is hampered by the combination of software unavailability, lack of customizability, and non-disparagement agreements (gag clauses) [3]. A need exists for an open-source EMR system designed for research.

Existing open-source EMR applications, like VistA [4] and OpenMRS [5], are free and unrestricted by non-disparagement agreements but are still challenging to customize because they implement a range of EMR core functions. Core functions—such as result and order management, communication, clinical decision and patient support, and administrative processes and reporting [6]—increase the size and complexity of the application’s codebase.

Many of these core functions of full-fledged EMR systems are not essential to investigators researching specific topics. For example, an EMR system’s result and order management views are sufficient to study patient data access patterns in the EMR (information-seeking behaviors [7, 8]), user experience (cognitive load, fatigue, acceptance [9, 10]), and task completion (time to task completion, accuracy of task completion, unintended consequences [2]). A lean EMR system designed for rapid customization and use by investigators will fill an important research need.

We report on the development, functionality, and implementation of a simple EMR system, useful for exploring user interface designs and evaluations in laboratory settings. As an open-source tool built from widely used open-source components, this software package provides a starting point for several types of EMR studies and, with further enhancements, could be extended to satisfy additional types of research.

## 2 Objective

To accelerate research in the design and usability of EMR systems, we developed a Python package that implements an EMR system designed for research. We describe its key features—including details of the user interface, how to add patient records, and its use in research—and suggest ways to contribute to this open-source package.

## 3 Methods

We implemented an EMR system designed for displaying patient data in laboratory-based research studies. The EMR system, called Simple EMR System, is built in Python using Bitnami Django [11]. System components—including a MySQL patient database, a user interface, and experimental business logic—follow a model view controller (MVC) architecture (see Figure 1).

**Figure 1:**
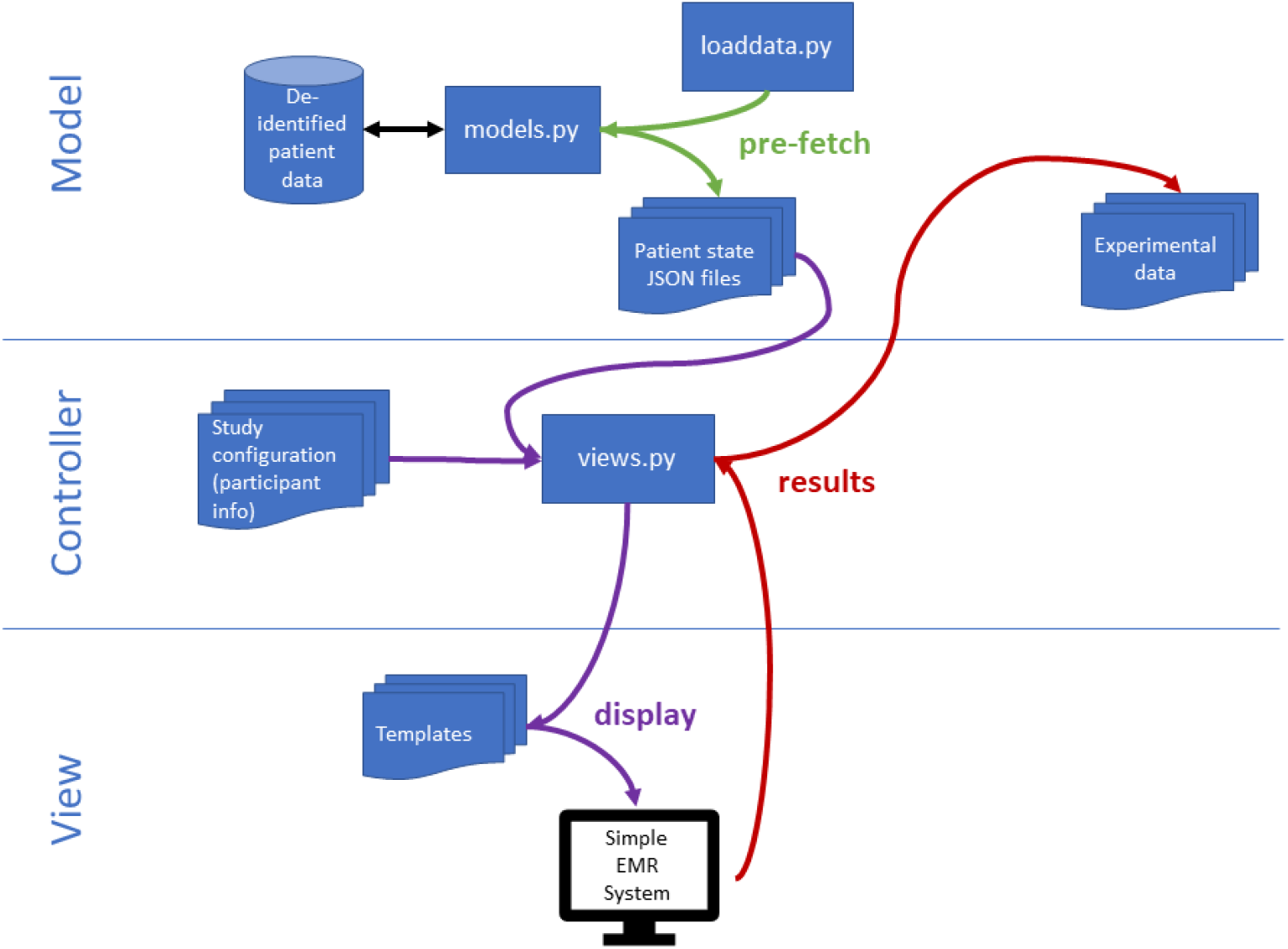
Simple EMR Systems’ MVC architecture. Patient data are pre-fetched and transformed to reduce load times. The case order and experimental conditions are managed in the controller via a configuration file for each user. The layout of the user interface is dictated by templates and accompanying JavaScript code. Experimental results, such as time to task completion or manual annotations of information-seeking behavior, are stored in text files.

MVC is a widely-used software design pattern that provides a framework for implementing a client-server application [12]. The **model** is a representation of the data that allows the data to be drawn from the underlying database without having to handle the complexities of creating, reading, updating, and deleting data in the database. To efficiently serve large amounts of patient data to the client, we implemented a pre-fetch function that queries each patient’s record up until a specified time point, transforms it into a JSON data structure, and stores it in a file. When a patient’s record is requested at runtime, the appropriate JSON file is loaded and immediately served to the client, resulting in faster load times.

The **view** is the presentation layer and constitutes the information that is viewed by the user in the interface of the Simple EMR System. Using the Django template language, JavaScript, and various libraries (Bootstrap, jQuery, Highstock), the HTML template pages are the access point for customizing the interface layout.

The **controller** manages the flow of information between the model and the view. It decides what information is obtained from the database via the model and sent to the view, as well as what information is obtained from the user via the view and relayed to the model. In the Simple EMR System, the controller is a combination of built-in Django functions and python scripts implementing business logic for determining what content to serve to the client. For example, in a research study that simulates clinical tasks across a series of patient cases, the case order and other simulation parameters are stored in supporting files that the controlling python script reads to serve the correct information to the participating clinician.

## 4 Results

### 4.1 Simple EMR User interface

The Simple EMR System presents patient data temporally in four scrollable columns (see Figure 2). From left to right, the first panel contains vital signs, ventilator settings, and intake and output, the second panel contains medication administrations, the third panel contains laboratory test results, and the fourth panel contains clinical notes and reports. The user interface includes interactive features to make using the temporal representation tenable (see Figure 3). For example, users adjust the displayed time range by click-and-dragging start and end point markers on a timeline. The time adjustments update all the temporal data plots to keep all displayed data in sync. The plots have blue and peach regions to indicate the normal range (y-axis) and the most recent 24 hours (x-axis), respectively. When an individual measurement is clicked, a vertical dotted line is placed at that time point on all plots. Further, the value of measurement appears in a pop-up box when the mouse is placed on a data point.

**Figure 2:**
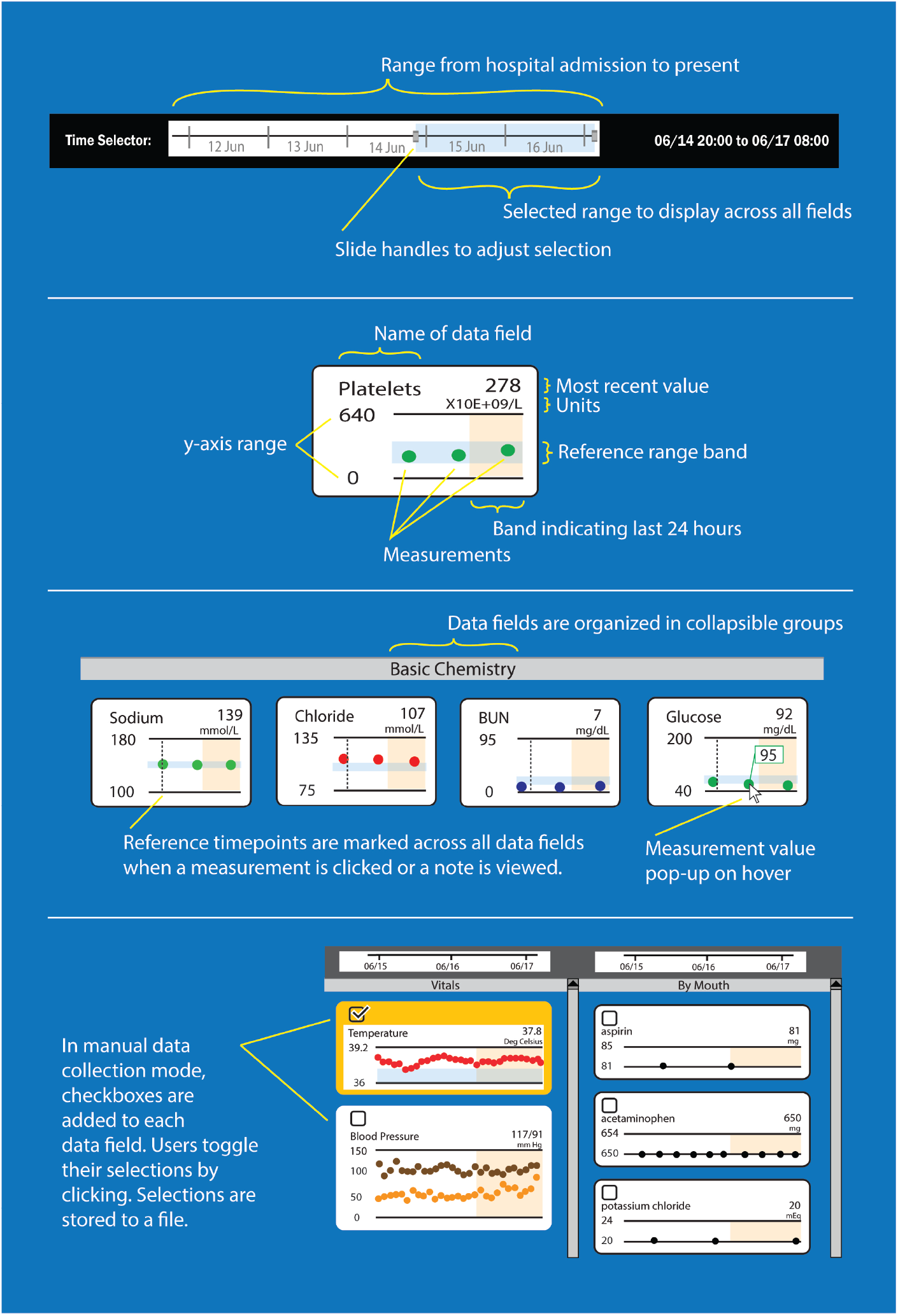
A screenshot of the user interface of the Simple EMR System. Most patient data are shown using temporal plots. At the top in black the interface shows patient demographics and a time selector, which is used for adjusting the display range for the temporal data plots. The data plots are organized into four scrollable columns. The left-most region shows vital signs, ventilator settings and fluid intake and output. The next region shows medication orders, organized by medication route. The next region shows laboratory test results, organized by panels (e.g., basic chemistry). On the data plots, the range of the x-axis is determined by the blue-selected region of the time selector at the top of the window. The final, right-most region shows clinical notes and reports, organized by type (consultant, progress, operation, radiology, electrocardiogram, microbiology, procedures, and others). At the lower right-hand region is an instruction and navigation region that can be customized for research.

**Figure 3:**
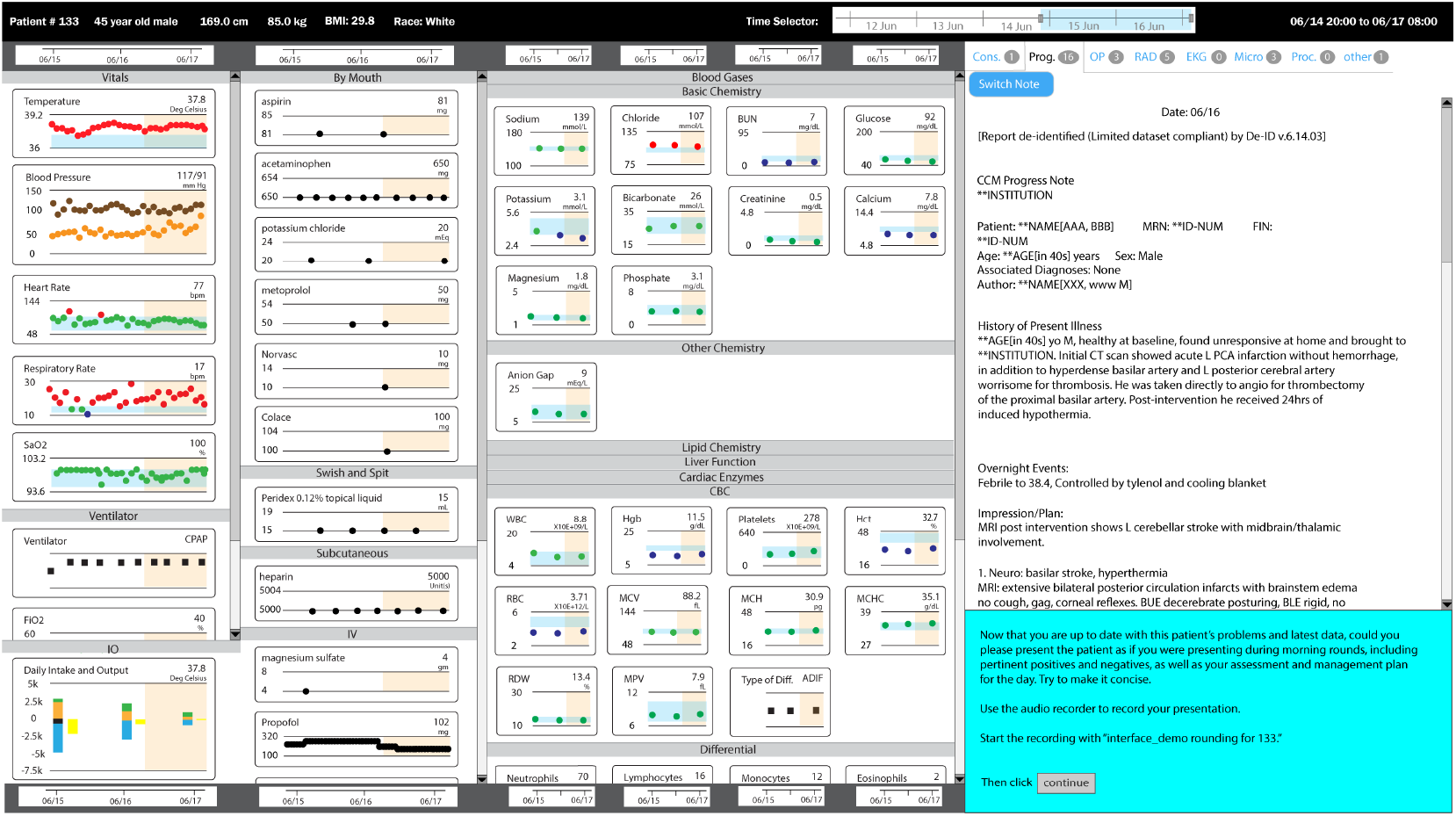
Details of some of the features of the user interface of the Simple EMR System. The first panel shows the adjustable time range that applies to all temporal plots. The second panel shows details of a single temporal data element such as platelet count. The third panel shows that laboratory tests results are organized into panels that are clinically meaningful, that reference timepoints can be added to aid users when interpreting data across multiple columns, and that individual measurements pop-up on cursor hover. The fourth panel shows data fields with check boxes that are used to indicate selections.

The temporal design, layout, and interactive features were influenced by feedback from critical care physicians across multiple iterations of prototypes and research studies.

The user interface has features that enable data collection for research. For example, study participants on reviewing patient cases can select relevant patient data by toggling a checkbox associated with a data element (see Figure 3). Also, the lower right-hand region of the user interface provides instruction and navigation areas that can be customized for research (see Figure 2). The system can be integrated with a low-cost eye-tracking device, as we have done, that is mounted at the bottom edge of the computer monitor that logs on-screen eye-gaze data.

The Simple EMR System software package with accompanying documentation is freely available on GitHub at https://github.com/ajk77/SimpleEMRSystem.

### 4.2 Patient records

The Simple EMR System includes three synthetic patient cases created by scrambling the contents of de-identified records from a larger set of patients. We include these synthetic cases to demonstrate the functionality of the system and to make system use concrete. The patient data are stored in JSON format in the resources folder.

Additional patient records can be loaded by either: (1) directly creating new JSON files using the same structure as the included synthetic patient records, or (2) connecting the system to a patient record database (e.g., MIMIC [13]) and executing a patient data loading script. This script extracts and transforms the raw data into a format that is readily consumed by the JavaScript libraries that generate the display, thus reducing load times. The script must be customized when connecting to different database schemas.

### 4.3 Example research use

The initial use case for the Simple EMR System was in support of the Learning EMR project where our goal was to build a data-driven method for prioritizing patient information in the EMR [14, 15, 16]. The project developed machine learning models to highlight patient data that are predicted to be relevant in the context of a specific clinical task [17]. The predictive models were trained on the information-seeking behavior of physicians when searching for relevant patient data in the EMR.

We used the Simple EMR System to collect clinician information-seeking behavior in two distinct ways: manually and with eye-tracking. In manual data collection, a checkbox is placed next to the plot for each data field. The user toggles the checkbox to select/deselect data fields of interest, and the system stores the list of selected data fields in a text file.

For eye-tracking, a low-cost eye-tracking device was mounted at the bottom of the computer monitor (see Figure 4). The Simple EMR System can be paired with a Tobii Eye Tracker 4C, to capture both a stream of eye-gaze coordinates and a stream of position coordinates for what is displayed onscreen. An algorithm then automatically maps eye-gaze to onscreen data elements to generate a list of what the user was viewing [18, 19, 20]. The scripts and accompanying documentation for incorporating eye-tracking with the Simple EMR System are freely available on GitHub at https://github.com/ajk77/EyeBrowserPy.

**Figure 4:**
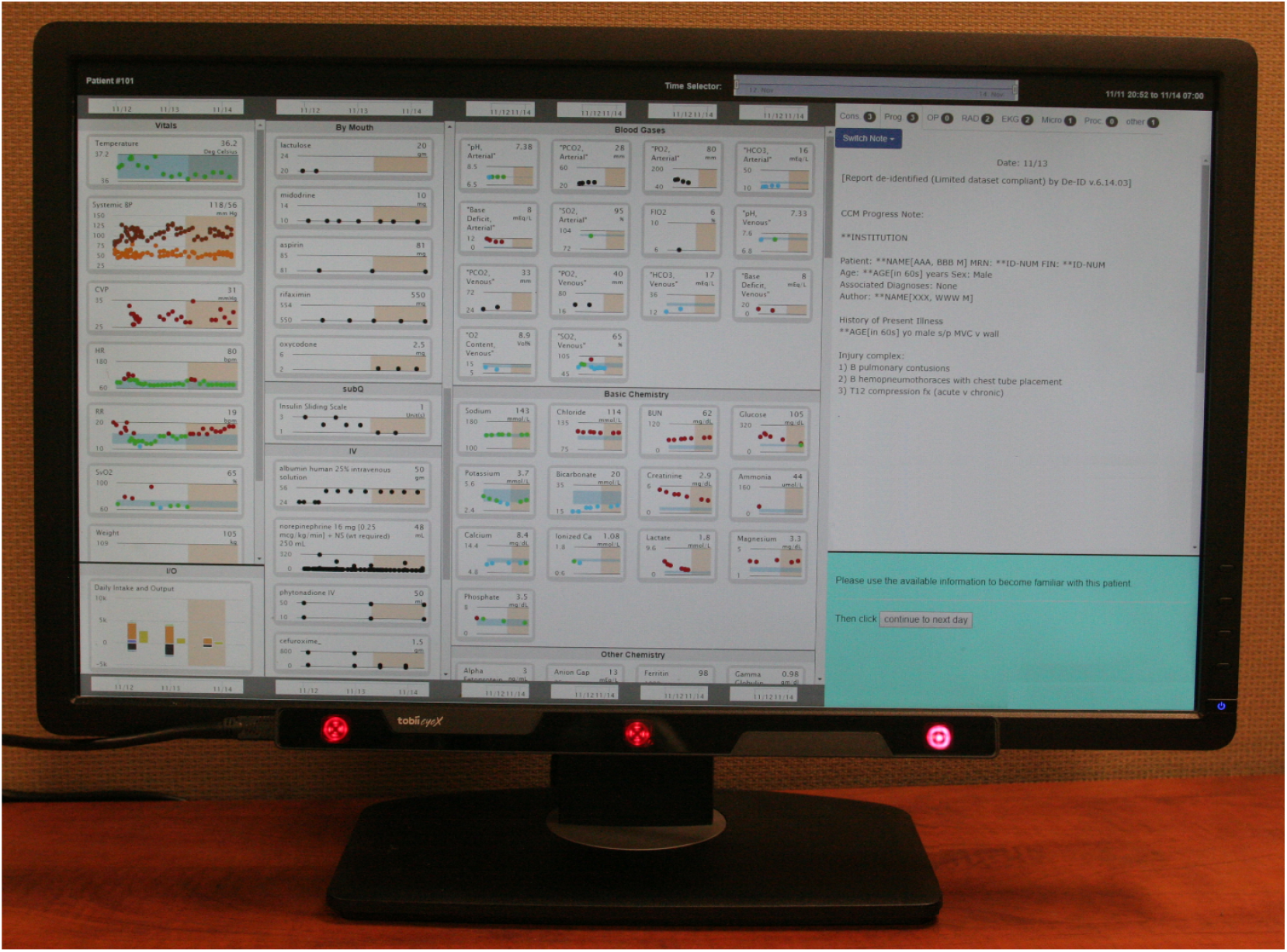
A low cost eye-tracking device is attached to the bottom of a computer monitor using a magnetic strip. The computer monitor is showing the interface of the Simple EMR System.

As part of the same work, we also used the Simple EMR System to test different means of prioritizing patient information, including selectively highlighting relevant patient data by changing the background color of some data elements.

## 5 Discussion

We developed an open-source EMR system designed for research. The Simple EMR System fills a gap for investigators who need a readily customizable EMR interface. It is well suited to support research about clinician information-seeking behaviors and adaptive user interfaces in terms measures that include task accuracy, time to completion, and cognitive load. Django’s MVC framework provides several advantages to investigators. The view provided by the Django template language is easy to modify and quickly make changes to the elements displayed on the screen. The model can be bypassed altogether if someone wishes to create patient cases in JSON format directly, rather than connecting to a database and customizing the data loading script. Finally, the python script that controls experimental parameters and case flow can also be modified independently from either the view or model.

The Simple EMR System is limited to the display functions of an EMR system. Although this limitation precludes it from being used in research involving order entry or other more complex EMR activities, the simple code base makes adoption and modifications easier. Compared to OpenMRS and VistA, the Simple EMR System is easier to deploy and customize for different experimental needs. Rapid and iterative deployment can keep research projects moving and enables A/B testing of many different interface layouts.

The Simple EMR System does not interact with vendor EMR systems that are currently in use in healthcare settings. Our goal in developing this system was not to replicate an entire modern EMR but to offer a simple and useful system for displaying patient data that mimics a real EMR in important ways to support research in laboratory settings. However, the open-source implementation provides a path toward integration with vendor system, as the python scripts we provide for loading of patient data might be modified to access vendor FHIR interfaces. Similarly, adaptation of the Simple EMR System to work within the SMART-on-FHIR framework is an interesting direction for future work [21].

We welcome investigators to contribute to the Simple EMR System. It is available under the GNU General Public License v3.0. We list potential avenues for improvement in Table 1.

**Table 1:**
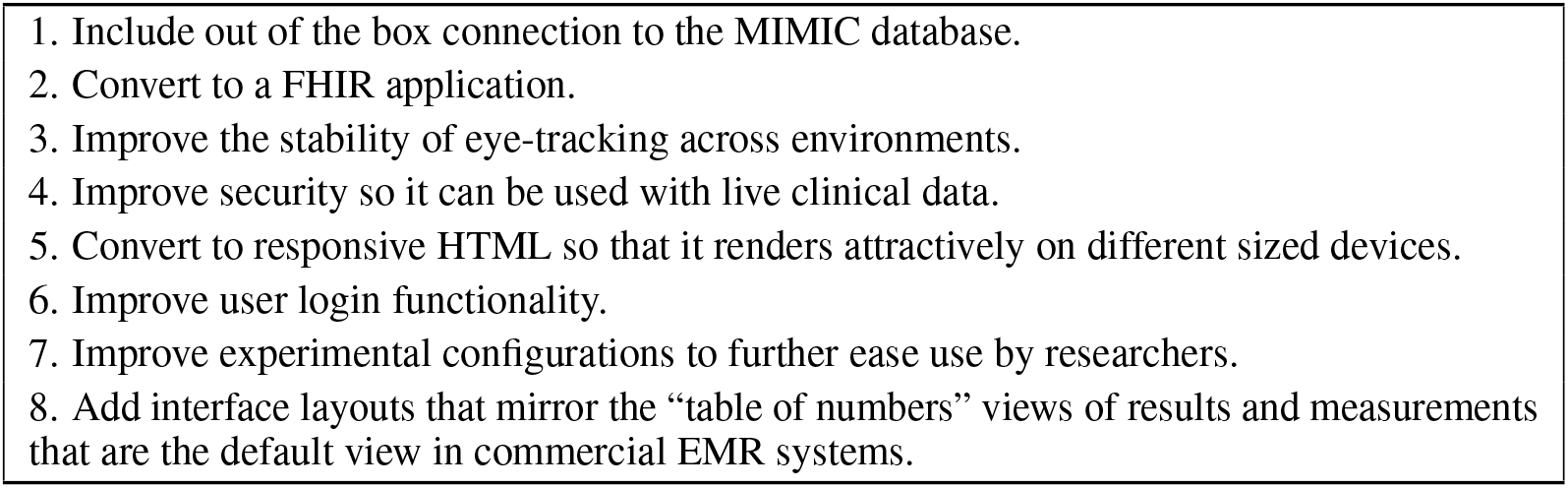
Community Opportunities for Contributing to the Simple EMR System.

## 6 Conclusion

We developed a Simple EMR System and have made it available to the research community. It provides a platform for EMR human factors research that can save researchers time by allowing for custom EMR scenarios and interface layouts. Beyond accelerating our understanding of clinician perception and preferences when viewing patient data, the system may lead to new ways of presenting patient data that reduce the cognitive burden and alleviate some causes of burnout. We welcome contributors to this open-source project.

## Data Availability

The Simple EMR System software package with accompanying documentation is freely available on GitHub at https://github.com/ajk77/SimpleEMRSystem. Eye-tracking for use with the Simple EMR System software is freely available on GitHub at https://github.com/ajk77/EyeBrowserPy

## Acknowledgments

The research reported in this paper was supported by the National Library of Medicine of the National Institutes of Health under award numbers T15 LM007059 and R01 LM012095. The content is solely the responsibility of the authors and does not necessarily represent the official views of the National Institutes of Health.

